# Emergency department revisits at thirty days are modestly explained by caregiver burden: a prospective cohort study

**DOI:** 10.1101/2024.09.25.24314385

**Authors:** Nathalie Germain, Annie Toulouse-Fournier, Rawane Samb, Émilie Côté, Vanessa Couture, Stéphane Turcotte, Michèle Morin, Yves Couturier, Lucas B. Chartier, Nadia Sourial, Samir Sinha, Don Melady, Marie-Soleil Hardy, Richard Fleet, France Légaré, Denis Roy, Holly O Witteman, Éric Mercier, Josée Chouinard, Josée Rivard, Marie-Josée Sirois, Joanie Robitaille, Patrick M. Archambault, LEARNING WISDOM investigators, Network of Canadian Emergency Researchers

## Abstract

**Importance:** Caregivers play a protective role in emergency department (ED) care transitions. When the demands of caregiving result in caregiver burden, ED returns can ensue.

**Objective:** We developed models describing how caregiver burden may predict ED revisits and admissions up to thirty days after discharge.

**Design:** This prospective cohort study nested within the LEARNING WISDOM clinical trial included older adults and their caregivers who underwent a transition of care from one of four EDs in Québec, Canada between January 1st, 2019, and December 21st, 2021.

**Setting:** This study occurred within an integrated health multi-site organization consisting of four acute care hospitals.

**Participants:** Patients aged 65 years or older who were discharged back to the community from the ED observation unit after being triaged to a stretcher on their index visit.

**Exposure:** Caregiver burden, as collected using the brief twelve-item Quebec French version of the Zarit Brief Burden Interview (ZBI).

**Main Outcomes and Measure:** Revisits to the ED were defined as a return to any ED in the 4-hospital network within 3, 7, or 30 days of the index visit. Admissions were return visits to the ED within 30 days resulting in hospitalization.

**Results:** Among 1,409 caregiver-patient dyads, ZBI scores averaged 7.33 (SD = 7.11). Most caregivers were women (69%). Caregivers were most often spouses (48%) of patients or children of patients (38%). Among all patients, 5.3% returned to the ED within 3 days, 9.4% returned within 7 days, 20.7% returned within 30 days and 6.2% were admitted within 30 days. Each point increase on the ZBI scale was associated with a 2.8% increase in the odds of a 30-day revisit to the ED (p = 0.03), but not in models with shorter time windows, nor for admissions. ZBI scores on 30-day ED revisits were moderated by the COVID-19 pandemic waves: the first inter-wave period attenuated the association.

**Conclusions and Relevance:** Caregiver burden may modestly predict ED revisits over 30 days. Future studies may enhance the management of ED revisits by predicting the longitudinal impact of caregiver burden on ED use in older adults.

**Trial Registration:** https://clinicaltrials.gov/ct2/show/NCT04093245

**Key points:** **Question:** Can caregiver burden predict emergency department (ED) revisits and admissions within 30 days of discharge among older adults?

**Findings:** In this prospective cohort study, higher caregiver burden was associated with a modest increase in the likelihood of 30-day ED revisits, though not with shorter-term revisits or admissions.

**Meaning:** Reducing caregiver burden may help prevent returns to the ED within 30 days among community-dwelling older adults.

## Background

Older adults are frequent users of emergency department (ED) services^1,2^. ED return visits after discharge from an index ED visit by older adult patients is a substantial contributing factor to ED overutilization^3^. Frequent ED use for complex needs is considered suboptimal, especially among older adults and indicates that those needs have not been adequately addressed^4–7^. Older adults are vulnerable to adverse outcomes related to ED visits, due in part to poor care transitions, declines in functional autonomy, lack of social support once discharged from the ED, comorbidities, and polypharmacy^8^. Both ED utilization and resource use intensity (e.g., diagnostic testing, consultation, length of stay) appear to increase with age^8^. Informal or family caregivers are often called upon to support this transition^9,10^.

Caregivers protect the health of those in their care^11^ and are also often included in care recipient assessments and decision-making^12^. As part of their role in patient care, they may endure physical, emotional, social, and financial strain, known collectively as caregiver burden^13,14^. Caregiver burden is associated with higher patient mortality^15^ and with admissions^16^. However, there is a knowledge gap in understanding if caregiver burden operationalized with questionnaires can predict increased ED revisits shortly after discharge. Our objective was to develop a model exploring the prognostic power of caregiver burden to explain unplanned ED revisits up to thirty days after discharge.

## Method

### Study design and context

This prospective cohort study was nested within the LEARNING WISDOM longitudinal cohort study^17^. We report our findings with the TRIPOD^18^ and STROBE^19^ guidelines.

The protocol for this study was approved by the *Centre intégré de santé et de services sociaux - Chaudière Appalaches* (CISSS-CA, Québec, Canada) Ethics Review Committee (project #2018-462, 2018-007). The LEARNING WISDOM cohort included older adults and their caregivers who underwent a transition of care following a visit to one of the four EDs within the CISSS-CA between January 1st, 2019, and December 21st, 2021. The CISSS-CA is an integrated health organization consisting of four acute care hospitals: Hôtel-Dieu de Lévis (HDL); a university teaching hospital), Hôpital de Saint-Georges (HSG), Hôpital de Montmagny (HDM), and Hôpital de Thetford Mines (HTM).

### Participants

The LEARNING WISDOM cohort included consenting patients aged at least 65 years, who had been discharged back to the community from the ED after being triaged to an observation unit stretcher on their index visit. Patients only seen in the ambulatory care section of the ED, admitted to hospital, transferred to another hospital, or transferred to a long-term care center following the index visit were excluded. Caregivers of older patients were informal non compensated caregivers, usually family members or friends, who provided support and assistance to patients in this cohort. Patients and their caregivers were required to understand French.

### Data collection

As part of a continuous quality improvement project led by the CISSS-CA, patients were contacted by telephone between 24 hours and up to seven days after ED discharge^20^. Patients were then invited to participate in a more in-depth research interview in the following days, and both patients and their caregivers were required to summarize—in their own words— their understanding of the study, based on the Nova Scotia Criteria during this second call to demonstrate informed consent^21^. After patients participated, they were asked if they consented to have their caregivers contacted by the research team. We conducted a structured interview to obtain demographic characteristics, followed by administering the Québec French version of the 12-item Zarit Brief Burden Interview (ZBI) to all participating caregivers^22^.

### Measures

We extracted hospital administrative data from MedGPS and MedUrge (MédiaMed Technologies, Mont-Saint-Hilaire, Québec, Canada) databases. Demographic and questionnaire data were collected using REDCap (Research Electronic Data Capture, Vanderbilt University, TN, USA) by trained research professionals^23,24^.

The Zarit Burden Interview (ZBI) is the most widely used instrument measuring caregiver burden with internal consistency indices ranging between 0.7 and 0.9^25–27^. In the twelve-item ZBI, questions include items about strain in the caregiver’s role and personal life associated with caregiving. Each question is scored by frequency in a five-point Likert scale (0 to 4), and scores are summed with higher scores indicating a higher degree of burden (range: 0– 48) (See Appendix A).

Outcome variables include patient revisits to the ED and hospital admissions on revisit. Revisits to the ED were defined as whether a given patient returned to any ED in the 4-hospital network within 3, 7 or 30 days of the index visit for any reason. Admissions include returns to the ED occurring within 30 days after the index visit that result in admission to the hospital. Index visits are defined as the patient’s first visit to the ED that required triage to a stretcher in the observation unit. Revisit intervals are associated with different outcomes. Early revisits within 3 to 7 days are generally considered failures of care coordination, while at 30 days failures are due to multifactorial factors of the care transition^28–32^.

Covariates included both patient and caregiver characteristics collected by trained research personnel over the telephone. For patients, we collected age, sex, income, education level, living situation (home with family, home alone, living in a care or retirement home), access to a family doctor, access to an appointment with a family doctor in a reasonable delay, access to transport and precedent visits to the ED over the past year. We also collected patient comorbidities using the Charlson Comorbidity Index (CCI)^33^, whether patients arrived by their own means or arrived by ambulance, the Canadian Triage and Acuity Scale (CTAS)^34^, and the time spent on a stretcher at the ED. For the CCI, we removed points allocated according to the age of the patient to consider age as an independent predictor variable. Caregiver characteristics of interest included their age, sex, ethnicity, income, education level, housing, and the nature of the caregiver-care recipient relationship (spouse, child-parent, other).

We also included the wave of COVID-19 pandemic according to Québec public health authorities at the time of the index visit. Data aggregation was performed to maximize the distinguishability of each stratum. Data grouping decisions can be found in Appendix B.

### Power analyses

We performed a-priori analyses to determine the estimated power to detect effects of interest (Appendix C). In simulations using normally distributed ZBI scores, 700 patients were sufficient to achieve statistical power (80%), and we estimated models could accommodate a maximum of 3 covariates and 3 interaction terms with ZBI scores as the predictor variable^35,36^.

### Logistic regression analyses

Using logistic regression modeling with a purposeful selection algorithm^37,38^ (Appendix D), we analyzed if caregiver burden among caregivers of older adult patients statistically explained ED revisits and ED revisits resulting in admissions^39,40^. All available clinically relevant data (Appendix B) were used in the development of the models. We also analyzed whether the COVID-19 pandemic period had moderated the relations between caregiver burden, revisits, and admissions. Data cleaning and analyses were conducted in R (version 4.3.0).

### Sensitivity and exploratory analyses

ZBI scores may have been biased by the timing of measurement. In some cases, due to delays in data collection, caregivers may have responded to the ZBI after the revisit to the ED may have occurred. We tested whether the coefficients, significance levels, and model fit statistics differed between dyads whose ZBI scores were collected before and those collected after the revisit occurred.

### Results Participants

The total LEARNING WISDOM cohort included 5,016 participants (Figure 1). Among these participants of the larger study, 1,819 allowed the research team to contact their caregiver, and 410 caregivers were excluded (6 unable to provide informed consent, 161 declined to or withdrew their participation, and 243 could not be reached), leaving 1,409 patient-caregiver dyads.

**Figure 1.**
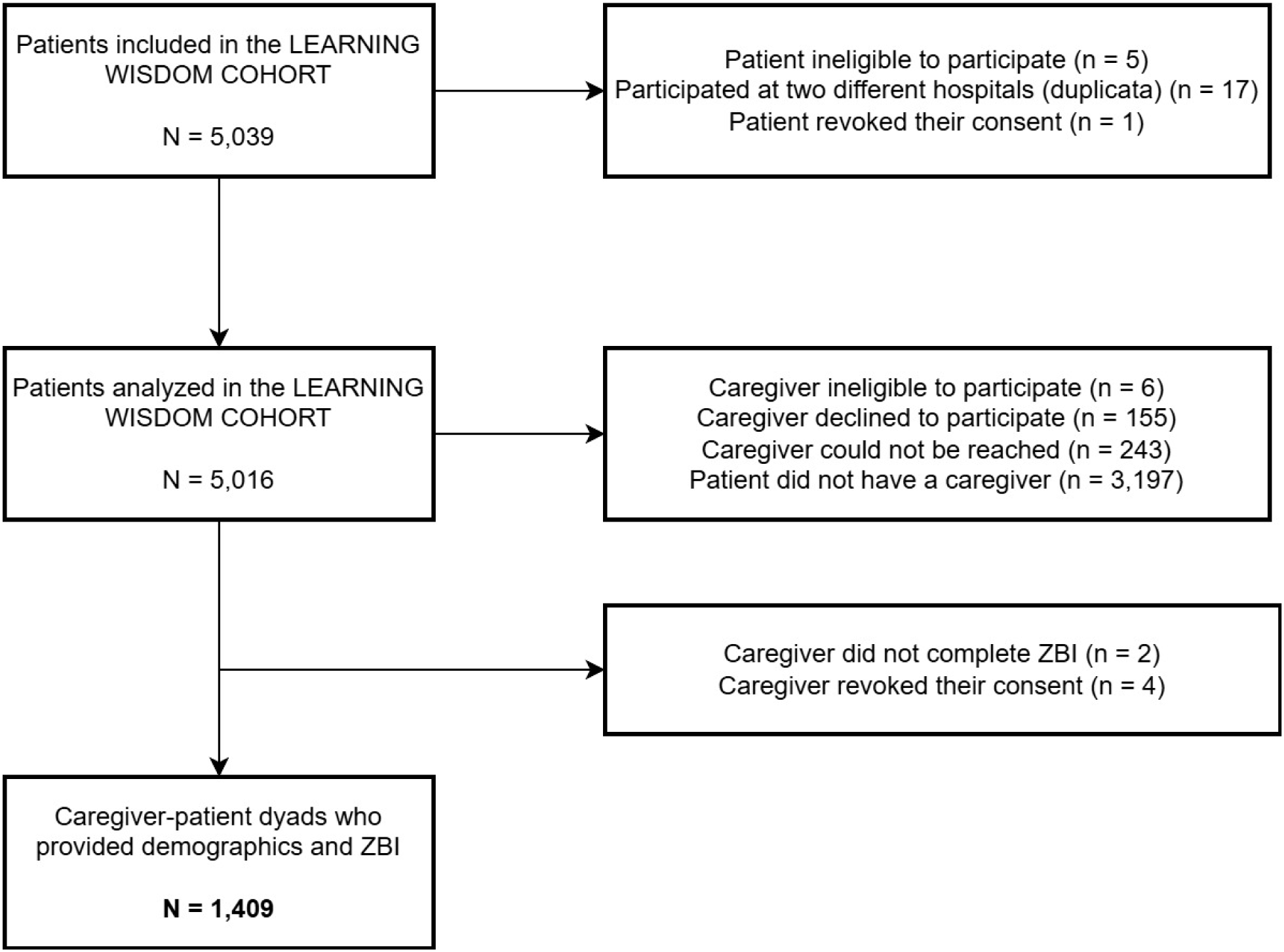
Flowchart describing the recruitment of patients and their caregivers.

ZBI scores averaged 7.33 (SD = 7.11). The internal consistency of ZBI scores was high (Cronbach’s alpha = .87, 95% CI = [.86, .89]). Among patients, 49.5% were women and 50.5% men. Among caregivers, women constituted the majority at 69.6%, contrasting with 30.4% men. Regarding caregiver-patient relationships, the largest proportion consisted of parent-child relationships at 48.0%, followed by spouses at 37.9%, and other family members or friends at 14.1%. Among all patients, 20.7% returned to the ED within 30 days of the index visit, 9.4% revisited within one week, and 5.3% within 72 hours, and 6.2% experienced a revisit resulting in admission within 30 days. Demographic characteristics stratified by those who revisited the ED within 30 days are found in Table 1.

**Table 1.** Demographic characteristics of patients and their caregivers.

|  | ED revisit<br>within 30 days<br>(N=292) | No ED revisit<br>within 30 days<br>(N=1117) |
| --- | --- | --- |
| **Patient sex** |  |  |
| Woman | 151 (51.7%) | 547 (49.0%) |
| Man | 141 (48.3%) | 570 (51.0%) |
| **Patient age** |  |  |
| Mean (SD) | 76.7 (7.29) | 77.2 (7.42) |
| Median [Min, Max] | 76.0 [65.0,<br>102] | 76.0 [65.0, 100] |
| **Arrival method on index visit** |  |  |
| Ambulance | 146 (50.0%) | 604 (54.1%) |
| Ambulant | 146 (50.0%) | 513 (45.9%) |
| **CTAS triage score on index visit<sup>\$</sup>** | | |
| 5 | 34 (11.6%) | 148 (13.2%) |
| 4 | 132 (45.2%) | 541 (48.4%) |
| 3 | 109 (37.3%) | 386 (34.6%) |
| 2 | 17 (5.8%) | 42 (3.8%) |
| **Hospital visited on index visit** |  |  |
| Hôtel-Dieu de Lévis (HDL) | 60 (20.5%) | 227 (20.3%) |
| Montmagny (HDM) | 98 (33.6%) | 449 (40.2%) |
| St-Georges-de-Beauce (HSG) | 56 (19.2%) | 203 (18.2%) |
| Thetford Mines (HTM) | 78 (26.7%) | 238 (21.3%) |
| **Time on stretcher at the index visit (hours)** |  |  |
| Mean (SD) | 11.1 (8.71) | 11.6 (8.60) |
| Median [Min, Max] | 8.39 [0.110,<br>52.4] | 8.72 [0, 70.2] |
| **Triage delay at the index visit**<br>**(hours)** |  |  |
| Mean (SD) | 0.0705 (0.155) | 0.0649 (0.152) |
| Median [Min, Max] | 0 [0, 1.20] | 0 [0, 1.32] |
| **Number of visits to the ED in the**<br>**last year** |  |  |
| Mean (SD) | 1.71 (2.19) | 1.16 (1.62) |
| Median [Min, Max] | 1.00 [0, 16.0] | 1.00 [0, 12.0] |
| **ED revisit**<br>**within 30 days** |  |  |
| ED revisit within 30 days | 292 (100%) | 0 (0%) |
| No ED revisit within 30 days | 0 (0%) | 1117 (100%) |
| **Social support (number of**<br>**persons in social circle, self-**<br>**report)** |  |  |
| Mean (SD) | 4.05 (4.41) | 4.11 (5.57) |
| Median [Min, Max] | 3.00 [0, 60.0] | 3.00 [0, 150] |
| **COVID-19 period when the**<br>**index ED visit occurred<sup>¶</sup>** |  |  |
| Pre-pandemic | 114 (39.0%) | 484 (43.3%) |
| Wave 1 | 41 (14.0%) | 158 (14.1%) |
| Between Wave 1 and Wave 2 | 16 (5.5%) | 60 (5.4%) |
| Wave 2 | 76 (26.0%) | 244 (21.8%) |
| Between Wave 2 and Wave 3 | 23 (7.9%) | 96 (8.6%) |
| Wave 3 | 22 (7.5%) | 75 (6.7%) |
| **Charlson Comorbidity Index** |  |  |
| Mean (SD) | 5.61 (2.37) | 5.35 (2.23) |
| Median [Min, Max] | 5.00 [2.00,<br>14.0] | 5.00 [2.00, 14.0] |
| **Caregiver sex** |  |  |
| Woman | 91 (31.2%) | 338 (30.3%) |
| Man | 201 (68.8%) | 779 (69.7%) |
| **Caregiver age** |  |  |
| Mean (SD) | 63.3 (12.0) | 64.0 (12.0) |
| Median [Min, Max] | 65.0 [28.0,<br>89.0] | 66.0 [24.0, 92.0] |
| **Patient has a family physician** |  |  |
| Yes | 16 (5.5%) | 81 (7.3%) |
| No | 276 (94.5%) | 1036 (92.7%) |
| **Patient can quickly consult their family physician** |  |  |
| Yes | 138 (47.3%) | 477 (42.7%) |
| No | 154 (52.7%) | 640 (57.3%) |
| **Patient has access to transport for medical care** |  |  |
| Yes | 18 (6.2%) | 52 (4.7%) |
| No | 274 (93.8%) | 1065 (95.3%) |
| **Patient education level** |  |  |
| Primary school | 141 (48.3%) | 534 (47.8%) |
| Secondary school | 116 (39.7%) | 464 (41.5%) |
| University | 35 (12.0%) | 119 (10.7%) |
| **Caregiver education level** |  |  |
| Primary school | 68 (23.3%) | 212 (19.0%) |
| Secondary school | 183 (62.7%) | 736 (65.9%) |
| University | 41 (14.0%) | 169 (15.1%) |
| **Caregiver-Patient relation** |  |  |
| Other family | 42 (14.4%) | 156 (14.0%) |
| Spouse | 116 (39.7%) | 418 (37.4%) |
| Parent-Child | 134 (45.9%) | 543 (48.6%) |
| **Patient annual income (\$ CAD) <sup>†</sup>** | | |
| No response | 128 (43.8%) | 438 (39.2%) |
| < 30,000\$ | 94 (32.2%) | 374 (33.5%) |
| > or equal to 30,000\$ | 70 (24.0%) | 305 (27.3%) |
| **Caregiver annual income (\$ CAD)** | | |
| No response | 130 (44.5%) | 467 (41.8%) |
| < 50,000\$ | 92 (31.5%) | 342 (30.6%) |
| > or equal to 50,000\$ | 70 (24.0%) | 308 (27.6%) |
| **Patient residence type** |  |  |
| Home | 48 (16.4%) | 174 (15.6%) |
| Home, alone | 179 (61.3%) | 725 (64.9%) |
| Care home | 65 (22.3%) | 218 (19.5%) |
| **Caregiver residence type** |  |  |
| Home | 7 (2.4%) | 17 (1.5%) |
| Home, alone | 239 (81.8%) | 952 (85.2%) |
| Care home | 46 (15.8%) | 148 (13.2%) |
| **ZBI Score** |  |  |
| Mean (SD) | 8.55 (7.42) | 7.01 (6.99) |
| Median [Min, Max] | 7.00 [0, 40.0] | 5.00 [0, 44.0] |

† Canadian dollars. § CTAS of 1: requires immediate care, 2: requires emergent care and rapid medical intervention, CTAS of 3: requires urgent care, CTAS of 4: requires less-urgent care, CTAS of 5: requires non-urgent care. ¶ Waves correspond to the Institut national de santé publique du Québec (INSPQ) definitions of the COVID-19 timeline in Québec: before the COVID-19 pandemic (01/01/2019 to 12/03/2020) and throughout waves one (13/03/2020 to 11/07/2020), two (23/08/2020 to 20/03/2021), three (21/03/2021 to 17/07/2021), four (18/07/2021 to 4/12/2021) and five (5/12/2021 to 12/03/2022).

### Models of 30-day revisits

The logistic regression model that best explained 30-day revisits to the ED included ZBI scores, ED visits in the preceding year, and the COVID-19 period. The COVID-19 period did not have a statistically significant main effect but did interact with ZBI scores. Although not statistically significant, it also interacted with ED visits in the past year (Table 2). Each added point on the ZBI scale was associated with a 2.84% increase in the odds of an early 30-day ED revisit, while controlling for covariates. Similarly, each ED visit in the past year was associated with a 11.9% increase in the odds of an early 30-day ED revisit. However, the second COVID-19 pandemic wave appeared to attenuate the association between ZBI scores and 30-day revisits, and the third wave appeared to attenuate the association between precedent ED visits and 30-day revisits. The model (Akaike information criterion (AIC) = 1424.6) was a good fit of the data (Goodness of Fit Test, X^2^(8) = 9.11, p = 0.332). The C-statistic, representing the model’s discriminative capabilities was low (C = 0.63, SE = 0.02).

**Table 2.** Model characteristics of logistic regression model explaining 30-day ED revisits.

| Characteristic | Univariate<br>OR<sup>†</sup> | Multivariate<br>OR | 95% CI<sup>‡</sup> | p-value |
| --- | --- | --- | --- | --- |
| **ZBI Score** | 1.03 | 1.03 | 1.00, 1.05 | **0.030** |
| **Previous ED visits** | 1.17 | 1.12 | 1.01, 1.24 | **0.028** |
| **Covid-19 Period** |  |  |  |  |
| *Pre-pandemic* | — | — | — |  |
| *Wave 1* | 1.10 | 0.61 | 0.28, 1.23 | 0.2 |
| *Between Wave 1 and Wave 2* | 1.13 | 2.27 | 0.89, 5.52 | **0.074** |
| *Wave 2* | 1.32 | 1.34 | 0.78, 2.30 | 0.3 |
| *Between Wave 2 and Wave 3* | 1.02 | 0.63 | 0.23, 1.55 | 0.3 |
| *Wave 3* | 1.25 | 1.56 | 0.67, 3.45 | 0.3 |
| **ZBI Score \* Covid-19 Period** |  |  |  |  |
| *ZBI Score \* Wave 1* | — | 1.04 | 0.98, 1.10 | 0.2 |
| *ZBI Score \** | — | 0.89 | 0.78, 0.97 | **0.027** |
| *Between Wave 1 and Wave 2* |  |  |  |  |
| *ZBI Score \* Wave 2* | — | 1.01 | 0.96, 1.05 | 0.8 |
| *ZBI Score \** | — | 1.03 | 0.95, 1.11 | 0.5 |
| *Between Wave 2 and Wave 3* |  |  |  |  |
| *ZBI Score \* Wave 3* | — | 0.96 | 0.89, 1.03 | 0.3 |
| **Previous ED visits \* Covid-19 Period** |  |  |  |  |
| *Previous ED visits \* Wave 1* | — | 1.23 | 0.99, 1.55 | **0.065** |
| *Previous ED visits \** | — | 1.14 | 0.80, 1.61 | 0.4 |
| *Between Wave 1 and Wave 2* |  |  |  |  |
| *Previous ED visits \* Wave 2* | — | 0.97 | 0.81, 1.15 | 0.7 |
| *Previous ED visits \** | — | 1.20 | 0.92, 1.61 | 0.2 |
| *Between Wave 2 and Wave 3* |  |  |  |  |
| *Previous ED visits \* Wave 3* | — | 1.02 | 0.80, 1.30 | 0.9 |

<sup>1</sup>OR = Odds Ratio, CI = Confidence Interval

Appendix E presents detailed model output for the models presented next. Figure 3 presents the ROC curves associated with each model.

**Figure 2.**
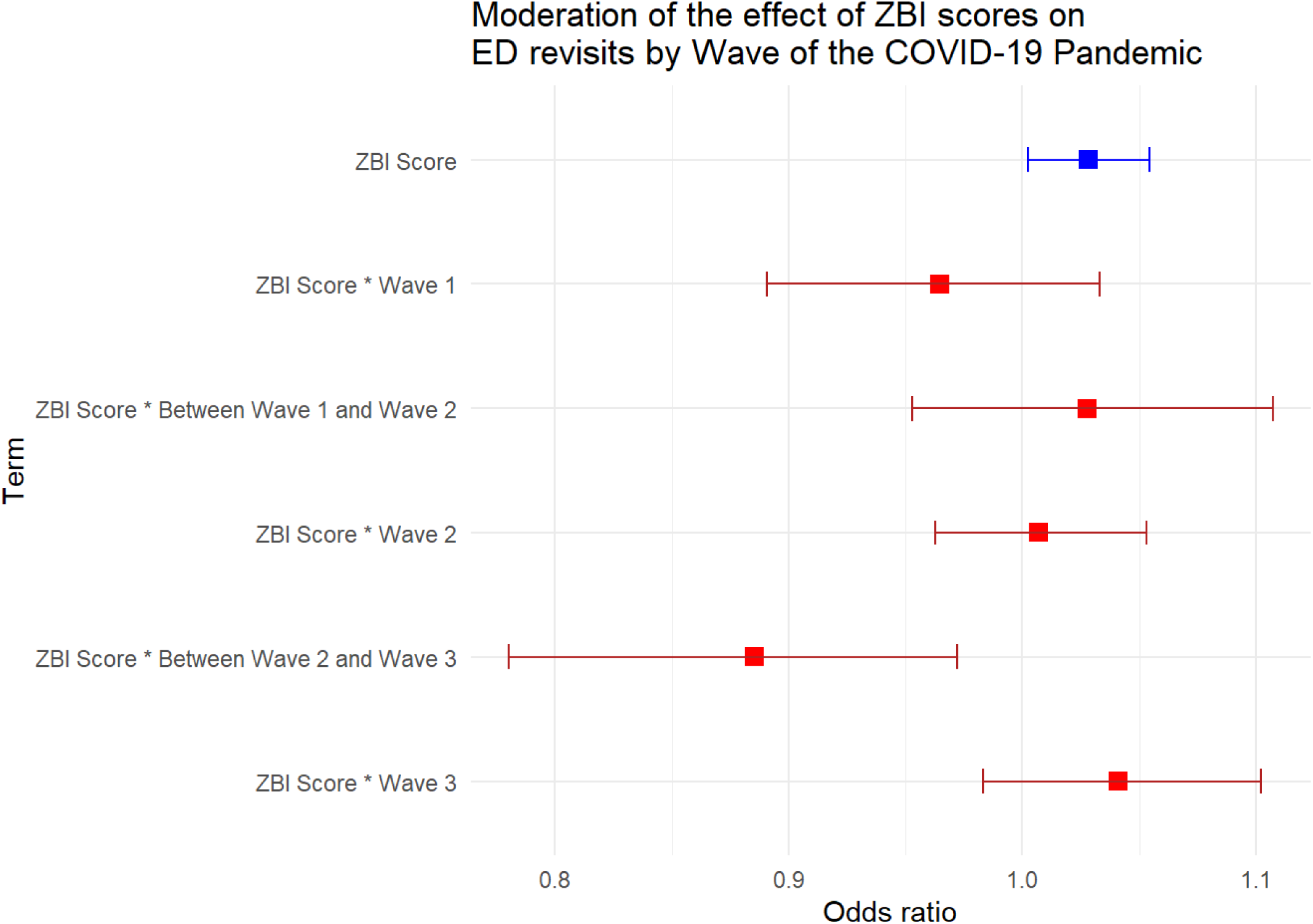
Moderation effect of ZBI scores on ED revisits by Wave of the COVID-19 pandemic. *Figure 2. Odds ratios and 95% confidence intervals for the interaction between ZBI scores and COVID-19 periods on thirty-day ED revisits. Red points represent the interaction effects of ZBI scores with over pandemic waves, while the blue point depicts the main effect of ZBI scores*.

**Figure 3.**
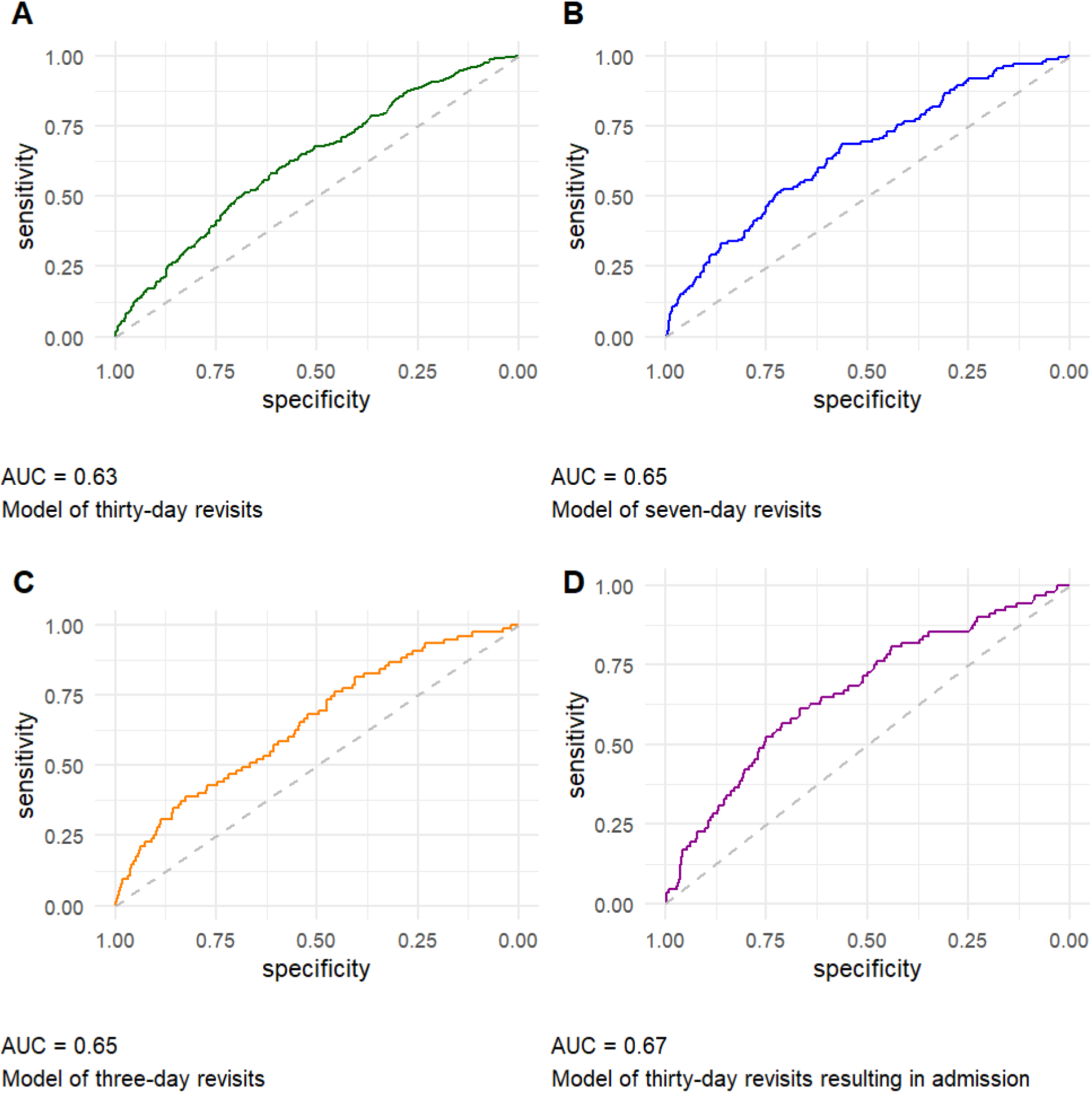
Receiver operating characteristics (ROC) curves associated with each logistic regression model. *Figure 3. A. Predictors of thirty-day revisits include ZBI score, number of ED visits in the previous year, and the COVID-19 period, which interacted with both ZBI score and past ED visits. B. Predictors of seven-day revisits include number of ED visits in the previous year, female sex, patient living in a care home, a caregiver living alone, and a CTAS triage score below 5. C. Predictors of three-day revisits include number of ED visits in the previous year, a caregiver living alone, a CTAS triage score below 5, and time on stretcher at the ED. D. Predictors of thirty-day revisits resulting in admission include Charlson score, a walk-in arrival to the ED, and a greater caregiver income*.

### Models of 7-day revisits

Female sex, ED visits in the pr) and year, a CTAS triage level of two at the index visit, patients living alone (p = .077), and having a caregiver residing home alone positively predicted revisits at 7 days whereas ED length of stay on stretcher (p = .059) was negatively related to the probability of an ED revisit. ZBI scores were not statistically significantly associated with the probability of a 7-day ED revisit (p = .55). There were no statistically significant interactions between these variables, nor the ZBI, nor the effect of the COVID-19 pandemic waves. The model was a good fit of the data (AIC = 865.6; Goodness of Fit Test, X^2^(8) = 4.33, p = .826). The C-statistic was low (C = 0.65 SE = 0.02).

### Models of 3-day revisits

ED visits in the last year, and a CTAS triage level of 4, 3, or 2 positively predicted revisits within 72 hours of the index visit. ED length of stay on a stretcher at the index visit and having a caregiver living alone (and not with the patient in their care), or in a care or a retirement home instead of a house or an apartment, were all negatively related to an ED revisit within 72 hours. ZBI scores were not significantly associated with 72-hour revisits (p = .68). There were no statistically significant interactions between these variables, nor the ZBI, nor the effect of the COVID-19 pandemic waves. The model was a good fit of the data (AIC = 580.64; Goodness of Fit Test, X^2^(8) = 2.93, p = .938). The C-statistic was low (C = 0.65 SE = 0.03).

### Models of 30-day admissions

A walk-in arrival at the index visit, and comorbidity index were statistically significantly associated with an ED revisit at 30 days resulting in a hospital admission. A higher annual caregiver revenue was protective against 30-day admissions. ZBI scores were not significantly associated with 30-day admissions (p = .24). There were no significant interactions. The model was a good fit of the data (AIC = 642.78, Goodness of Fit Test, X^2^(8) = 8.49, p = .386). The C-statistic was low (C = 0.67, SE = 0.03).

### Sensitivity analyses

Most caregivers had their data collected before the patient revisited to the ED (N = 1099, 78%). However, 310 patients (22%) revisited the ED before their caregiver was recruited. To determine if this affected the association between ZBI scores and ED revisits at 30 days, we split the dataset in two groups (caregiver burden measured before ED revisit and caregiver burden measured after ED revisit) and re-performed the first model. The coefficient for ZBI scores did not change significantly between these two models (OR = 1.022) and was very similar to the model containing all 1409 dyads (OR = 1.028). However, the C-statistic for the model containing ZBI data collected before patients returned to the ED (C = 0.68, SE = 0.03) was greater than the model containing patients who returned to the ED before the ZBI was collected (C = 0.61, SE = 0.03). When the ZBI was collected before the revisit, predictive ability in 30-day revisits was improved, but only the effects associated with pandemic waves changed (Appendix F).

## Discussion

We analyzed the association between caregiver burden of care and ED revisits, and subsequent hospital admissions within thirty days, among a large cohort of older community-dwelling adults. We adjusted for factors related to both caregiver burden and the tendency for repeat ED usage. ZBI scores were significantly associated with revisits at 30 days, holding important covariates constant. Other authors report that caregiver burden, when treated as a continuous variable, is associated with ED use among patients with major neurocognitive disorders, but its effect size was also small as was the case in our study^41^.

The effect of ZBI scores and precedent visits on 30-day ED revisits was moderated by the COVID-19 pandemic waves, with the first inter-wave period attenuating the association, likely because older adults were encouraged to distance themselves from healthcare services^42,43^. The models for revisits within 72 hours and 7 days identified gender, living conditions, and prior ED visits as factors that influenced the likelihood of a revisit, but ZBI scores were not a significant predictor in these shorter-term revisit models. Factors positively associated with 30-day admissions after an ED visit included a walk-in arrival at the ED index visit and higher scores on the age-adjusted Charlson Comorbidity Index, whereas higher annual caregiver revenue was protective against such admissions. We interpret these findings as evidence that caregiver burden may contribute to a negative care transition, which is associated with 30-day revisits, whereas shorter intervals and admissions related closer to pre-existing reliance on the ED and comorbidities.

The modest area under the curve (C-statistic) for all regression models suggest there are important missing variables at play, which might include the chronicity or acuteness of the presentation reason at the ED (e.g., chronic heart failure versus myocardial infarction), the frailty of patients and caregivers, and the health professional seen in the ED prior to discharge (e.g., consulting with a specialized geriatric emergency medicine nurse or a geriatrician). This also suggests a heterogeneous sample of patients presenting to the ED, but as the ED is the front door to the health system, some heterogeneity in sampling is expected. Caregiver burden is also known to be a highly personal and intersectional experience, which poses additional challenges to heterogeneity^44^.

In most cases, we were able to collect the ZBI before patients revisited the ED. While this presents a temporal bias, there is evidence to suggest that caregiver burden is stable at 5 months,^45^ 6 to 12 months,^46^ and 1 to 2 years^47^ after initial measurement, respectively. We had previously found that caregiver burden for caregivers in this cohort already experiencing some burden increased slightly following the index visit of an ED care transition^48^. Further research on caregiver burden as a predictor of emergency department recidivism would benefit from a longitudinal design to assess burden levels at discharge and during follow-up to parse out if fluctuations in burden are short-term or indicative of long-term trends among caregivers. In practice at the ED, the addition of a long-form caregiver burden questionnaire may not be feasible, but there is an ultra-short version of the ZBI (the ZBI-1)25 which could be tested prospectively to verify if systematic screening of caregiver burden in the ED and mitigation strategies put in place prior to ED discharge could reduce ED revisits^25,49^. Some such strategies include involving caregivers as active members of the care team,^50^ and caregiver navigators who review and disseminate information about local support services throughout the care transition^51^. Caregiver burden itself can be lessened through improving caregiving self-efficacy and with social support services designed to assist with chores and errands^48,52,53^.

### Strengths and limitations

Strengths of our study include the inclusion of a large prospective cohort from both urban and rural communities, the use of psychometrically validated tools, and thorough regression fit testing and variable selection aimed at parsimony and alignment with theoretical frameworks. Limitations include some potential selection biases. We administered our questionnaires via phone calls, which may have prevented the participation of patients who hear less well, and we excluded patients with neurocognitive disorders as was required by the ethics committee to ensure informed and competent consent. Bias in responding may have arisen from patients and caregivers who may have responded to questionnaires in a socially desirable way. Caregivers of patients with neurocognitive disorders are known to have higher degrees of caregiver burden^54,55^. Most caregiver data was collected before the patient revisited the ED, and although ZBI data collection conducted before the revisit slightly improved the predictive ability of 30-day ED revisits, it did not significantly alter model coefficients.

## Conclusion

Among caregivers of community-dwelling older adults, caregiver burden was associated with an increased likelihood of ED revisits within 30 days, although not for shorter 3 and 7-day revisit intervals, and not for revisits resulting in admissions. Our models had only modest predictive ability, indicating potential missing variables and unaccounted heterogeneity in this population. Future studies may consider measuring caregiver burden at ED discharge and leveraging longitudinal designs to deepen the predictive capabilities of caregiver burden in relation to ED use. This may improve our understanding of caregiver burden and its management to prevent ED revisits in older community-dwelling adults.

## Funding and acknowledgements Funding

The LEARNING WISDOM clinical trial was funded by an Embedded Clinician Salary Award (ECRA) awarded to PMA from the Canadian Institutes for Health Research (CIHR) (#201603), a Fonds de recherche du Québec – Santé (FRQS) Senior Clinical Scholar Award (#283211), and a CIHR Project Grant (#378616). Work on this article was supported by a Master’s Award: Canada Graduate Scholarships Award (CIHR) awarded to NG (#202112). The funding bodies had no role in the design of the study, collection, or analysis of the data, interpretation of the results, or writing of the manuscript. The authors do not have any conflicts of interest to declare.

## Data Availability

Analysis code is consultable in a public repository on GitHub. Please contact the corresponding author for a link to the repository. Anonymized data are available from the corresponding author on reasonable request.

## Acknowledgements

We acknowledge the invaluable support of participating patients and their caregivers. We also thank Lise Lavoie, David Buckeridge, Audrey-Anne Brousseau, Clémence Dallaire, Annie Leblanc, Marcel Émond, Isabelle Pelletier and Jean-Louis Denis for their support and expertise in planning and contributing to the LEARNING WISDOM project. Finally, we thank professors Aida Eslami and André Tourigny of Université Laval for their in-depth review of both this article and all the scholarly output that went into this project.

## Network of Emergency Researchers Patrick M. Archambault and Marcel Émond Data availability

Analysis code is consultable in a public GitHub repository. Anonymized data are available from the corresponding author on reasonable request.

## References

1. Aminzadeh F, Dalziel WB. Older adults in the emergency department: A systematic review of patterns of use, adverse outcomes, and effectiveness of interventions. Annals of Emergency Medicine. 2002;39(3):238–247. doi:10.1067/mem.2002.121523

2. Ukkonen M, Jämsen E, Zeitlin R, Pauniaho SL. Emergency department visits in older patients: A population-based survey. BMC emergency medicine. 2019;19(1):20. doi:10.1186/s12873-019-0236-3

3. Sheikh S. Risk Factors Associated with Emergency Department Recidivism in the Older Adult. Western Journal of Emergency Medicine. 2019;20(6):931. doi:10.5811/westjem.2019.7.43073

4. Chiu Y, Courteau J, Dufour I, Vanasse A, Hudon C. Machine learning to improve frequent emergency department use prediction: A retrospective cohort study. Scientific Reports. 2023;13(1). doi:10.1038/s41598-023-27568-6

5. Gettel CJ, Voils CI, Bristol AA, et al. Care transitions and social needs: A Geriatric Emergency care Applied Research (GEAR) Network scoping review and consensus statement. Academic Emergency Medicine: Official Journal of the Society for Academic Emergency Medicine. 2021;28(12):1430–1439. doi:10.1111/acem.14360

6. Hunold KM, Richmond NL, Waller AE, Cutchin MP, Voss PR, Platts-Mills TF. Primary care availability and emergency department use by older adults: A population-based analysis. Journal of the American Geriatrics Society. 2014;62(9):1699–1706. doi:10.1111/jgs.12984

7. Jones A, Maclagan LC, Watt JA, et al. Reasons for repeated emergency department visits among community-dwelling older adults with dementia in Ontario, Canada. Journal of the American Geriatrics Society. 2022;70(6):1745–1753. doi:10.1111/jgs.17726

8. Latham LP, Ackroyd-Stolarz S. Emergency department utilization by older adults: A descriptive study. Canadian geriatrics journal: CGJ. 2014;17(4):118–125. doi:10.5770/cgj.17.108

9. Samaras N, Chevalley T, Samaras D, Gold G. Older patients in the emergency department: A review. Annals of Emergency Medicine. 2010;56(3):261–269. doi:10.1016/j.annemergmed.2010.04.015

10. Mitchell SE, Laurens V, Weigel GM, et al. Care Transitions From Patient and Caregiver Perspectives. Annals of Family Medicine. 2018;16(3):225–231. doi:10.1370/afm.2222

11. McCusker J, Cetin-Sahin D, Cossette S, et al. How Older Adults Experience an Emergency Department Visit: Development and Validation of Measures. Annals of Emergency Medicine. 2018;71(6):755–766.e4. doi:10.1016/j.annemergmed.2018.01.009

12. Sloan JP. Protocols in Primary Care Geriatrics. 2nd ed. Springer; 1997. Accessed October 17, 2024. 10.1007/978-1-4612-1884-5

13. Bonin-Guillaume S, Durand AC, Yahi F, et al. Predictive factors for early unplanned rehospitalization of older adults after an ED visit: Role of the caregiver burden. Aging Clinical and Experimental Research. 2015;27(6):883–891. doi:10.1007/s40520-015-0347-y

14. Zarit SH, Reever KE, Bach-Peterson J. Relatives of the Impaired Elderly: Correlates of Feelings of Burden1. The Gerontologist. 1980;20(6):649–655. doi:10.1093/geront/20.6.649

15. Schulz R, Beach SR, Friedman EM. Caregiving Factors as Predictors of Care Recipient Mortality. The American Journal of Geriatric Psychiatry: Official Journal of the American Association for Geriatric Psychiatry. 2021;29(3):295–303. doi:10.1016/j.jagp.2020.06.025

16. Afonso-Argilés FJ, Meyer G, Stephan A, et al. Emergency department and hospital admissions among people with dementia living at home or in nursing homes: Results of the European RightTimePlaceCare project on their frequency, associated factors and costs. BMC geriatrics. 2020;20(1):453. doi:10.1186/s12877-020-01835-x

17. Archambault PM, Rivard J, Smith PY, et al. Learning Integrated Health System to Mobilize Context-Adapted Knowledge With a Wiki Platform to Improve the Transitions of Frail Seniors From Hospitals and Emergency Departments to the Community (LEARNING WISDOM): Protocol for a Mixed-Methods Implementation Study. JMIR research protocols. 2020;9(8):e17363. doi:10.2196/17363

18. Collins GS, Reitsma JB, Altman DG, Moons KGM. Transparent reporting of a multivariable prediction model for individual prognosis or diagnosis (TRIPOD): The TRIPOD statement. BMJ (Clinical research ed). 2015;350:g7594. doi:10.1136/bmj.g7594

19. Elm E von, Altman DG, Egger M, et al. The strengthening the reporting of observational studies in epidemiology (STROBE) statement: Guidelines for reporting observational studies. Journal of Clinical Epidemiology. 2008;61(4):344–349. doi:10.1016/j.jclinepi.2007.11.008

20. Couture V, Germain N, Côté É, et al. Transitions of care for older adults discharged home from the emergency department: An inductive thematic content analysis of patient comments. BMC Geriatrics. 2024;24(1):8. doi:10.1186/s12877-023-04482-0

21. Wiebe E, Kelly M, McMorrow T, Tremblay-Huet S, Hennawy M. Assessment of capacity to give informed consent for medical assistance in dying: A qualitative study of clinicians’ experience. CMAJ Open. 2021;9(2):E358. doi:10.9778/cmajo.20200136

22. Hébert R, Bravo G, Préville M. Reliability, Validity and Reference Values of the Zarit Burden Interview for Assessing Informal Caregivers of Community-Dwelling Older Persons with Dementia. Canadian Journal on Aging / La Revue canadienne du vieillissement. 2000;19(4):494–507. doi:10.1017/S0714980800012484

23. Harris PA, Taylor R, Thielke R, Payne J, Gonzalez N, Conde JG. Research electronic data capture (REDCap)–a metadata-driven methodology and workflow process for providing translational research informatics support. Journal of Biomedical Informatics. 2009;42(2):377–381. doi:10.1016/j.jbi.2008.08.010

24. Harris PA, Taylor R, Minor BL, et al. The REDCap consortium: Building an international community of software platform partners. Journal of Biomedical Informatics. 2019;95:103208. doi:10.1016/j.jbi.2019.103208

25. Kühnel MB, Ramsenthaler C, Bausewein C, Fegg M, Hodiamont F. Validation of two short versions of the Zarit Burden Interview in the palliative care setting: A questionnaire to assess the burden of informal caregivers. Supportive Care in Cancer: Official Journal of the Multinational Association of Supportive Care in Cancer. 2020;28(11):5185–5193. doi:10.1007/s00520-019-05288-w

26. Bianchi M, Flesch LD, Alves EV da C, Batistoni SST, Neri AL. Zarit Burden Interview Psychometric Indicators Applied in Older People Caregivers of Other Elderly. Revista Latino-Americana De Enfermagem. 2016;24:e2835. doi:10.1590/1518-8345.1379.2835

27. Lu L, Wang L, Yang X, Feng Q. Zarit Caregiver Burden Interview: Development, reliability and validity of the Chinese version. Psychiatry and Clinical Neurosciences. 2009;63(6):730–734. doi:10.1111/j.1440-1819.2009.02019.x

28. Rising KL, Victor TW, Hollander JE, Carr BG. Patient returns to the emergency department: The time-to-return curve. Academic Emergency Medicine: Official Journal of the Society for Academic Emergency Medicine. 2014;21(8):864–871. doi:10.1111/acem.12442

29. LaMantia MA, Platts-Mills TF, Biese K, et al. Predicting hospital admission and returns to the emergency department for elderly patients. Academic Emergency Medicine: Official Journal of the Society for Academic Emergency Medicine. 2010;17(3):252–259. doi:10.1111/j.1553-2712.2009.00675.x

30. Hess EP, Brison RJ, Perry JJ, et al. Development of a clinical prediction rule for 30-day cardiac events in emergency department patients with chest pain and possible acute coronary syndrome. Annals of Emergency Medicine. 2012;59(2):115–125.e1. doi:10.1016/j.annemergmed.2011.07.026

31. Taupin D, Anderson TS, Merchant EA, et al. Preventability of 30-Day Hospital Revisits Following Admission with COVID-19 at an Academic Medical Center. Joint Commission Journal on Quality and Patient Safety. 2021;47(11):696. doi:10.1016/j.jcjq.2021.08.011

32. Müller J, Keller DI, Slankamenac K. Unplanned revisits of older patients to the emergency department. Frontiers in Disaster and Emergency Medicine. 2024;2. doi:10.3389/femer.2024.1342904

33. Charlson M, Szatrowski TP, Peterson J, Gold J. Validation of a combined comorbidity index. Journal of Clinical Epidemiology. 1994;47(11):1245–1251. doi:10.1016/0895-4356(94)90129-5

34. Beveridge R, Ducharme J, Janes L, Beaulieu S, Walter S. Reliability of the Canadian emergency department triage and acuity scale: Interrater agreement. Annals of Emergency Medicine. 1999;34(2):155–159. doi:10.1016/s0196-0644(99)70223-4

35. Demidenko E. Sample size determination for logistic regression revisited. Statistics in Medicine. 2007;26(18):3385–3397. doi:10.1002/sim.2771

36. Zhang Z. Practical Statistical Power Analysis Using WebPower and R. (Yuan KH, ed.). ISDSA Press; 2018.

37. Bursac Z, Gauss CH, Williams DK, Hosmer DW. Purposeful selection of variables in logistic regression. Source Code for Biology and Medicine. 2008;3(1):17. doi:10.1186/1751-0473-3-17

38. Hosmer DW, Lemeshow S, Sturdivant RX. Model-Building Strategies and Methods for Logistic Regression. In: *Applied Logistic Regression*. John Wiley & Sons, Ltd; 2013:89–151. doi:10.1002/9781118548387.ch4

39. Flynn Longmire CV, Knight BG. Confirmatory factor analysis of a brief version of the Zarit Burden Interview in Black and White dementia caregivers. The Gerontologist. 2011;51(4):453–462. doi:10.1093/geront/gnr011

40. Hagell P, Alvariza A, Westergren A, Årestedt K. Assessment of Burden Among Family Caregivers of People With Parkinson’s Disease Using the Zarit Burden Interview. Journal of Pain and Symptom Management. 2017;53(2):272–278. doi:10.1016/j.jpainsymman.2016.09.007

41. Guterman EL, Allen IE, Josephson SA, et al. Association Between Caregiver Depression and Emergency Department Use Among Patients With Dementia. JAMA neurology. 2019;76(10):1166–1173. doi:10.1001/jamaneurol.2019.1820

42. Khanassov V, Ilali M, Ruiz AS, Rojas-Rozo L, Sourial R. Telemedicine in primary care of older adults: A qualitative study. BMC Primary Care. 2024;25(1):259. doi:10.1186/s12875-024-02518-x

43. Briere J, Wang SH, Khanam UA, Lawson J, Goodridge D. Quality of life and well-being during the COVID-19 pandemic: Associations with loneliness and social isolation in a cross-sectional, online survey of 2,207 community-dwelling older Canadians. BMC Geriatrics. 2023;23(1):615. doi:10.1186/s12877-023-04350-x

44. Liu R, Chi I, Wu S. Caregiving Burden Among Caregivers of People With Dementia Through the Lens of Intersectionality. The Gerontologist. 2022;62(5):650–661. doi:10.1093/geront/gnab146

45. O’Hara RE, Hull JG, Lyons KD, et al. Impact on caregiver burden of a patient-focused palliative care intervention for patients with advanced cancer. Palliative & Supportive Care. 2010;8(4):395–404. doi:10.1017/S1478951510000258

46. Marsh NV, Kersel DA, Havill JA, Sleigh JW. Caregiver burden during the year following severe traumatic brain injury. Journal of Clinical and Experimental Neuropsychology. 2002;24(4):434–447. doi:10.1076/jcen.24.4.434.1030

47. Manskow US, Friborg O, Røe C, Braine M, Damsgard E, Anke A. Patterns of change and stability in caregiver burden and life satisfaction from 1 to 2 years after severe traumatic brain injury: A Norwegian longitudinal study. NeuroRehabilitation. 2017;40(2):211–222. doi:10.3233/NRE-161406

48. Germain N, Jémus-Gonzalez E, Couture V, et al. Caregivers’ burden of care during emergency department care transitions among older adults: A mixed methods cohort study. BMC geriatrics. 2024;24(1):788. doi:10.1186/s12877-024-05388-1

49. Stagg B, Larner AJ. Zarit Burden Interview: Pragmatic study in a dedicated cognitive function clinic. Progress in Neurology and Psychiatry. 2015;19(4):23–27. doi:10.1002/pnp.390

50. Tyagi S, Koh GCH, Luo N, et al. Role of caregiver factors in outpatient medical follow-up post-stroke: Observational study in Singapore. BMC Family Practice. 2021;22:74. doi:10.1186/s12875-021-01405-z

51. Reblin M, Ketcher D, McCormick R, et al. A randomized wait-list controlled trial of a social support intervention for caregivers of patients with primary malignant brain tumor. BMC health services research. 2021;21(1):360. doi:10.1186/s12913-021-06372-w

52. Leung DYP, Chan HYL, Chiu PKC, Lo RSK, Lee LLY. Source of Social Support and Caregiving Self-Efficacy on Caregiver Burden and Patient’s Quality of Life: A Path Analysis on Patients with Palliative Care Needs and Their Caregivers. International Journal of Environmental Research and Public Health. 2020;17(15):5457. doi:10.3390/ijerph17155457

53. Arlotto S, Gentile S, Blin A, Durand AC, Bonin-Guillaume S. Caregiver Burden Is Reduced by Social Support Services for Non-Dependent Elderly Persons: Pre-Post Study of 569 Caregivers. International Journal of Environmental Research and Public Health. 2022;19(20):13610. doi:10.3390/ijerph192013610

54. Brodaty H, Donkin M. Family caregivers of people with dementia. Dialogues in Clinical Neuroscience. 2009;11(2):217–228. doi:10.31887/DCNS.2009.11.2/hbrodaty

55. Dauphinot V, Ravier A, Novais T, et al. Relationship Between Comorbidities in Patients With Cognitive Complaint and Caregiver Burden: A Cross-Sectional Study. Journal of the American Medical Directors Association. 2016;17(3):232–237. doi:10.1016/j.jamda.2015.10.011

