## Supplemental Files: Appendices A to F for "Emergency department revisits at thirty days are modestly explained by caregiver burden: a prospective cohort study"

### Additional File 1. Appendix A.

### 1.TRIPOD Checklist

| **Section/Topic** |  | **Checklist Item** | **Page** |
| --- | --- | --- | --- |
| **Title and abstract** | | | |
| Title | 1 | Identify the study as developing and/or validating a multivariable prediction model, the target population, and the outcome to be predicted. | 1 |
| Abstract | 2 | Provide a summary of objectives, study design, setting, participants, sample size, predictors, outcome, statistical analysis, results, and conclusions. | 2 |
| **Introduction** | | | |
| Background and objectives | 3a | Explain the medical context (including whether diagnostic or prognostic) and rationale for developing or validating the multivariable prediction model, including references to existing models. | 3 |
|  | 3b | Specify the objectives, including whether the study describes the development or validation of the model or both. | 3-4 |
| **Methods** | | | |
| Source of data | 4a | Describe the study design or source of data (e.g., randomized trial, cohort, or registry data), separately for the development and validation data sets, if applicable. | 6 |
|  | 4b | Specify the key study dates, including start of accrual; end of accrual; and, if applicable, end of follow-up. | 5 |
| Participants | 5a | Specify key elements of the study setting (e.g., primary care, secondary care, general population) including number and location of centres. | 5 |
|  | 5b | Describe eligibility criteria for participants. | 5 |
|  | 5c | Give details of treatments received, if relevant. | 5 |
| Outcome | 6a | Clearly define the outcome that is predicted by the prediction model, including how and when assessed. | 6-7 |
|  | 6b | Report any actions to blind assessment of the outcome to be predicted. | N/A |
| Predictors | 7a | Clearly define all predictors used in developing or validating the multivariable prediction model, including how and when they were measured. | 6-7 |
|  | 7b | Report any actions to blind assessment of predictors for the outcome and other predictors. | N/A |
| Sample size | 8 | Explain how the study size was arrived at. | 7-8 |
| Missing data | 9 | Describe how missing data were handled (e.g., complete-case analysis, single imputation, multiple imputation) with details of any imputation method. | 6 |
| Statistical analysis methods | 10a | Describe how predictors were handled in the analyses. | 7-8 |
|  | 10b | Specify type of model, all model-building procedures (including any predictor selection), and method for internal validation. | 7-8-9 |
|  | 10d | Specify all measures used to assess model performance and, if relevant, to compare multiple models. | 8 |
| Risk groups | 11 | Provide details on how risk groups were created, if done. | N/A |
| **Results** | | | |
| Participants | 13a | Describe the flow of participants through the study, including the number of participants with and without the outcome and, if applicable, a summary of the follow-up time. A diagram may be helpful. | 9  Figure 1 |
|  | 13b | Describe the characteristics of the participants (basic demographics, clinical features, available predictors), including the number of participants with missing data for predictors and outcome. | 9 Table 1 |
| Model development | 14a | Specify the number of participants and outcome events in each analysis. | 9 Table 1 |
|  | 14b | If done, report the unadjusted association between each candidate predictor and outcome. | Appendix B |
| Model specification | 15a | Present the full prediction model to allow predictions for individuals (i.e., all regression coefficients, and model intercept or baseline survival at a given time point). | Table 2  Appendix B |
|  | 15b | Explain how to the use the prediction model. | N/A |
| Model performance | 16 | Report performance measures (with CIs) for the prediction model. | Figure 2  Pages 9-11 |
| **Discussion** | | | |
| Limitations | 18 | Discuss any limitations of the study (such as nonrepresentative sample, few events per predictor, missing data). | 12, 14 |
| Interpretation | 19b | Give an overall interpretation of the results, considering objectives, limitations, and results from similar studies, and other relevant evidence. | 12, 13 |
| Implications | 20 | Discuss the potential clinical use of the model and implications for future research. | 14 |
| **Other information** | | | |
| Supplementary information | 21 | Provide information about the availability of supplementary resources, such as study protocol, Web calculator, and data sets. | Appendices |
| Funding | 22 | Give the source of funding and the role of the funders for the present study. | Funding and acknowledgements |

### 2.STROBE Checklist

|  | Item No | Recommendation |
| --- | --- | --- |
| **Title and abstract** | 1 | (*a*) Indicate the study’s design with a commonly used term in the title or the abstract |
|  |  | (*b*) Provide in the abstract an informative and balanced summary of what was done and what was found |
| Introduction | | |
| Background/rationale | 2 | Explain the scientific background and rationale for the investigation being reported |
| Objectives | 3 | State specific objectives, including any prespecified hypotheses |
| Methods | | |
| Study design | 4 | Present key elements of study design early in the paper |
| Setting | 5 | Describe the setting, locations, and relevant dates, including periods of recruitment, exposure, follow-up, and data collection |
| Participants | 6 | (*a*) Give the eligibility criteria, and the sources and methods of selection of participants. Describe methods of follow-up |
|  |  | (*b*) For matched studies, give matching criteria and number of exposed and unexposed |
| Variables | 7 | Clearly define all outcomes, exposures, predictors, potential confounders, and effect modifiers. Give diagnostic criteria, if applicable |
| Data sources/ measurement | 8* | For each variable of interest, give sources of data and details of methods of assessment (measurement). Describe comparability of assessment methods if there is more than one group |
| Bias | 9 | Describe any efforts to address potential sources of bias |
| Study size | 10 | Explain how the study size was arrived at |
| Quantitative variables | 11 | Explain how quantitative variables were handled in the analyses. If applicable, describe which groupings were chosen and why |
| Statistical methods | 12 | (*a*) Describe all statistical methods, including those used to control for confounding |
|  |  | (*b*) Describe any methods used to examine subgroups and interactions |
|  |  | (*c*) Explain how missing data were addressed |
|  |  | (*d*) If applicable, explain how loss to follow-up was addressed |
|  |  | (*e*) Describe any sensitivity analyses |
| Results | | |
| Participants | 13* | (a) Report numbers of individuals at each stage of study—eg numbers potentially eligible, examined for eligibility, confirmed eligible, included in the study, completing follow-up, and analysed |
|  |  | (b) Give reasons for non-participation at each stage |
|  |  | (c) Consider use of a flow diagram |
| Descriptive data | 14* | (a) Give characteristics of study participants (eg demographic, clinical, social) and information on exposures and potential confounders |
|  |  | (b) Indicate number of participants with missing data for each variable of interest |
|  |  | (c) Summarise follow-up time (eg, average and total amount) |
| Outcome data | 15* | Report numbers of outcome events or summary measures over time |
| Main results | 16 | (*a*) Give unadjusted estimates and, if applicable, confounder-adjusted estimates and their precision (eg, 95% confidence interval). Make clear which confounders were adjusted for and why they were included |
|  |  | (*b*) Report category boundaries when continuous variables were categorized |
|  |  | (*c*) If relevant, consider translating estimates of relative risk into absolute risk for a meaningful time period |
| Other analyses | 17 | Report other analyses done—eg analyses of subgroups and interactions, and sensitivity analyses |
| Discussion | | |
| Key results | 18 | Summarise key results with reference to study objectives |
| Limitations | 19 | Discuss limitations of the study, taking into account sources of potential bias or imprecision. Discuss both direction and magnitude of any potential bias |
| Interpretation | 20 | Give a cautious overall interpretation of results considering objectives, limitations, multiplicity of analyses, results from similar studies, and other relevant evidence |
| Generalisability | 21 | Discuss the generalisability (external validity) of the study results |
| Other information | | |
| Funding | 22 | Give the source of funding and the role of the funders for the present study and, if applicable, for the original study on which the present article is based |

### Additional File 2. Appendix B. List of variables collected and their transformations

| **Categorical or factor variables** | | |
| --- | --- | --- |
| **Re-coded categorical variable** | **Re-coded levels**  **(Translated)** | **Original levels as described in the French questionnaire** |
| Patient education level | Primary school* | Études primaires |
|  | Secondary school | Études secondaires (DES)  Formation professionnelle (DEP, ASP)  Études collégiales (DEC) |
|  | University | Baccalauréat  Études de cycles supérieurs (2e ou 3e cycle) |
| Caregiver education level | Primary school* | Études primaires |
|  | Secondary school | Études secondaires (DES)  Formation professionnelle (DEP, ASP)  Études collégiales (DEC) |
|  | University | Baccalauréat  Études de cycles supérieurs (2e ou 3e cycle) |
| Caregiver-Patient relation | Other family member or friend* | Soeur/frère  Amie/Ami  Petite-fille/petit-fils  Nièce/neveu  Belle-filles/beau-fils  Autre |
|  | Spouse | Conjoint/Conjointe |
|  | Parent-Child | Fils/Fille |
| Patient annual income  ($ CAD) | < 30,000$ | Moins de 10 000$  10 000 à 19 999$  20 000 à 29 999$ |
|  | > or equal to 30,000$ | 30 000 à 39 999$  40 000 à 49 999$  50 000 à 59 999$  60 000 à 69 999$  70 000 à 79 999$  80 000 à 89 999$  90 000 à 99 999$  Plus de 100 000$ |
|  | No response* | Préfère ne pas répondre  Inconnu / manquant |
| Caregiver annual income  ($ CAD) | < 50,000$ | Moins de 10 000$  10 000 à 19 999$  20 000 à 29 999$  30 000 à 39 999$  40 000 à 49 999$ |
|  | > or equal to 50,000$ | 50 000 à 59 999$  60 000 à 69 999$  70 000 à 79 999$  80 000 à 89 999$  90 000 à 99 999$  Plus de 100 000$ |
|  | No response* | Préfère ne pas répondre  Inconnu / manquant |
| Patient residence type | Care home | Résidence privée pour personnes aînés avec présence infirmière 24h/24  Résidence privée pour personnes aînés sans infirmière sur place  Centre hospitalier de soins longue durée (CHSLD) |
|  | Home, with others* | Domicile, partagé |
|  | Home, alone | Domicile, seul  Ressources intermédiaires ou de type familial (RI ou RTF)  Habitation à loyer modique (HLM) |
| Caregiver residence type | Care home | Résidence privée pour personnes aînés avec présence infirmière 24h/24  Résidence privée pour personnes aînés sans infirmière sur place  Centre hospitalier de soins longue durée (CHSLD) |
|  | Home, with others* | Domicile, partagé |
|  | Home, alone | Domicile, seul  Ressources intermédiaires ou de type familial (RI ou RTF)  Habitation à loyer modique (HLM) |

* Reference level

| **Numeric variables** | | |
| --- | --- | --- |
| **Variable name** | **Definition** | **Calculation** |
| **ZBI score** | Score of ZBI questionnaire | Sum of all ZBI items |
| **Time on stretcher (hours)** | Time patient spent on stretcher at the ED | Calculated by MedGPS |
| **Triage delay (hours)** | Delay between arrival at the ED and triage | Calculated by MedGPS |
| **Visits to the ED in the last year** | Number of unique visits to the ED 365 days before the index visit | Count of visits between 365 days before index and the index visit |
| **Visits to the ED in following 30 days** | Number of unique visits to the ED 30 days after the index visit | Count of visits between the index and the 30 days after the index visit |
| **Charlson Comorbidity Index** | Predicts the ten-year mortality for a patient with comorbid conditions | Weighted sum of comorbidity item scores |
| **Charlson Comorbidity Index (without age)** | Indice of comorbidity burden in each patient | Weighted sum of comorbidity item scores with the age-item removed |
| **Revisit at 30 days** | Whether or not a 30-day revisit occurred | Identifies whether a revisit occurred in the time between the date and time the patient left the ED + 30 days |
| **Revisit at 7 days** | Whether or not a 7-day revisit occurred | Identifies whether a revisit occurred in the time between the date and time the patient left the ED + 7 days |
| **Revisit at 3 days** | Whether or not a 3-day revisit occurred | Identifies whether a revisit occurred in the time between the date and time the patient left the ED + 3 days |
| **Revisit resulting in admission at 30 days** | Whether or not a 30-day revisit occurred resulting in admission | Identifies whether a revisit occurred in the time between the date and time the patient left the ED + 30 days AND the revisit was classified as an admission |

### Additional File 3. Appendix C. A-priori power analysis

We performed a-priori analyses to determine the estimated power to detect effects of interest. First, we assumed a very small effect size (OR = 1.2) of a positive association between ZBI scores and the likelihood of returning to the ED within 30 days, with a statistical significance level of 0.05. We also assumed a prevalence of 15% for 30-day ED visits. These estimates were based on previous work using the ZBI to predict hospital readmissions [1] and previous studies on ED use and revisit rates in older adult populations [2–6]. Using methods described by Demidenko [7] and Zhang and Yuan [8], power curves were plotted using logistic regression simulations with an ED revisit as the dependent variable, and the ZBI score as the single predictor variable. In simulations using normally distributed and lognormal ZBI scores [9,10], 1100 and 700 patients were sufficient to achieve a statistical power of 80%, respectively (Figures A and B).

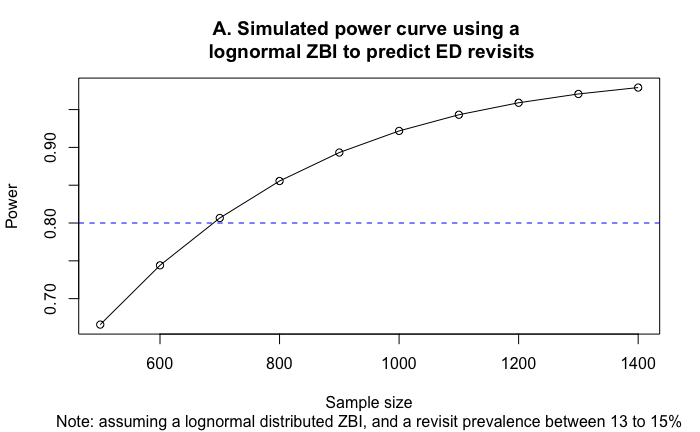

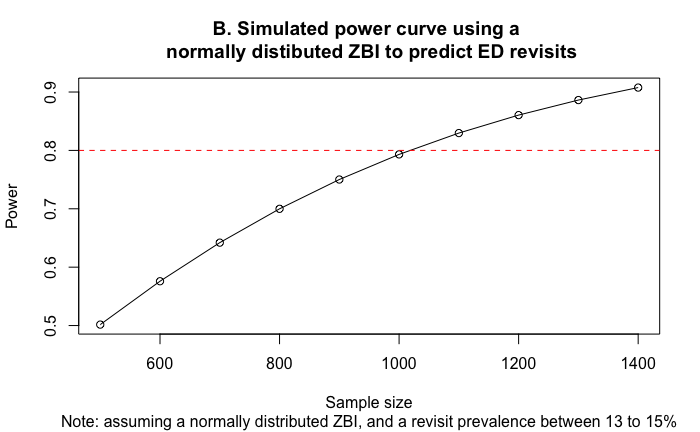

Simulations were then performed with the same parameters as described above to determine the number of covariates that could be accommodated while maintaining a statistical power of 80%. We had projected the number of caregiver-patient dyads to be between 1400 and 1500. For each successive logistic regression model assuming a normally distributed ZBI score, we added a covariate and an interaction effect between the ZBI scores and the covariate. Based on the results of these simulations, the model could accommodate a maximum of 3 covariates and 3 interaction terms with ZBI scores as the predictor variable (Figure C).

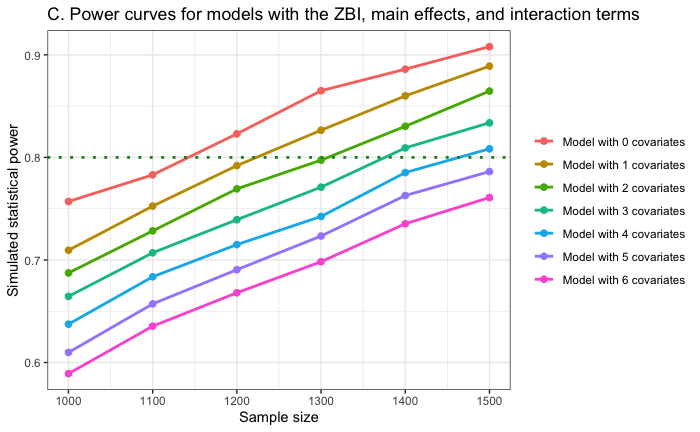

**References for Appendix C**

1. Fitriana I, Setiati S, Rizal EW *et al.* Malnutrition and depression as predictors for 30-day unplanned readmission in older patient: a prospective cohort study to develop 7-point scoring system. *BMC Geriatrics* 2021;**21**:256.

2. Gruneir A, Fung K, Fischer HD *et al.* Care setting and 30-day hospital readmissions among older adults: a population-based cohort study. *CMAJ* 2018;**190**:E1124–33.

3. Gruneir A, Cigsar C, Wang X *et al.* Repeat emergency department visits by nursing home residents: a cohort study using health administrative data. *BMC Geriatrics* 2018;**18**:157.

4. Simpson M, Sergi C, Malsch A *et al.* Association of Geriatric Emergency Department post-discharge referral order and follow-up with healthcare utilization. *J Am Geriatr Soc* 2023;**71**:821–31.

5. Hamilton MP, Bellolio F, Jeffery MM *et al.* Risk of falls is associated with 30-day mortality among older adults in the emergency department. *The American Journal of Emergency Medicine* 2024;**79**:122–6.

6. Sun BC, Burstin HR, Brennan TA. Predictors and Outcomes of Frequent Emergency Department Users. *Academic Emergency Medicine* 2003;**10**:320–8.

7. Demidenko E. Sample size determination for logistic regression revisited. *Statistics in Medicine* 2007;**26**:3385–97.

8. Zhang Z, Yuan K-H. *Practical Statistical Power Analysis Using Webpower and R*., 2018.

9. Flynn Longmire CV, Knight BG. Confirmatory factor analysis of a brief version of the Zarit Burden Interview in Black and White dementia caregivers. *Gerontologist* 2011;**51**:453–62.

10. Hagell P, Alvariza A, Westergren A *et al.* Assessment of Burden Among Family Caregivers of People With Parkinson’s Disease Using the Zarit Burden Interview. *J Pain Symptom Manage* 2017;**53**:272–8.

### Additional File 4. Appendix D. Purposeful selection procedure as outlined in Hosmer et al., 2013.

*Step 1* of this method involved conducting univariate tests of covariates on the outcome variable, selecting any variables with a *p-value* < .25. *Step 2* used all the covariates identified in *Step 1* to construct a base model. In *Step 3*, iterative variable selection was conducted, and non-significant covariates (*p-value* < .05) were removed unless they acted as confounders, identified by changes exceeding 20% in parameter estimates. In *Step 4*, any variables not selected in *Step 2* were added back into the model. If any of these variables showed statistically significant main effects, they were added back into the model, which was then our *preliminary main effects model*. In *Step 5*, the assumption of linearity was tested for each continuous variable. This assumption of linearity in the logit referred to the linear relation between the log odds of the outcome variable and the predictor variables. In *Step 6*, the main effects in the model were tested for interaction effects. Interaction between two covariates implied that the effect of each variable was not constant over levels of the other variable. Lastly, in *Step 7*, goodness of fit was assessed, along with an evaluation of the discriminatory power of this final model using a receiver operating characteristic (ROC) curve.

### Additional File 5. Appendix E. Model output for 7-day revisits, 3-day revisits and 30-day revisits resulting in an admission.

A. Model characteristics from logistic regression model on 7-day revisits

| **Characteristic** | | **Univariate OR**^1^ | **Multivariate OR**^1^ | **95% CI**^1^ | **p-value** |
| --- | --- | --- | --- | --- | --- |
| **ZBI Score** | | 1.01 | 1.01 | 0.98, 1.03 | 0.6 |
| **Previous ED visits** | | 1.14 | 1.15 | 1.05, 1.25 | **0.002** |
| **Time on stretcher at the ED** | | 0.98 | 0.98 | 0.95, 1.00 | **0.059** |
| **Patient sex** | |  |  |  |  |
| *Woman* | | — | — | — |  |
| *Man* | | 0.60 | 0.60 | 0.40, 0.88 | **0.010** |
| **Patient residence type** | |  |  |  |  |
| *Home* | | — | — | — |  |
| *Home, alone* | | 1.47 | 1.59 | 0.86, 3.15 | 0.2 |
| *Care home* | | 1.70 | 2.46 | 1.24, 5.14 | **0.012** |
| **Caregiver residence type** | |  |  |  |  |
| *Home* | | — | — | — |  |
| *Home, alone* | | 0.31 | 0.26 | 0.09, 0.80 | **0.013** |
| *Care home* | | 0.25 | 0.19 | 0.06, 0.63 | **0.005** |
| **Triage (CTAS) on index visit** | |  |  |  |  |
| *5* | | — | — | — |  |
| *4* | | 1.81 | 1.95 | 1.01, 4.13 | **0.060** |
| *3* | | 1.89 | 1.96 | 1.00, 4.22 | **0.064** |
| *2* | | 3.51 | 3.95 | 1.52, 10.3 | **0.004** |
|  | ^1^OR = Odds Ratio, CI = Confidence interval for multivariate OR | | | | |

B. Model characteristics from logistic regression model on 3-day revisits

| **Characteristic** | **Univariate OR**^1^ | **Multivariate OR**^1^ | **95% CI**^1^ | **p-value** |
| --- | --- | --- | --- | --- |
| **ZBI Score** | 1.01 | 1.01 | 0.97, 1.04 | 0.7 |
| **Previous ED visits** | 1.13 | 1.13 | 1.01, 1.25 | **0.023** |
| **Caregiver residence type** |  |  |  |  |
| *Home* | — | — | — |  |
| *Home, alone* | 0.27 | 0.30 | 0.11, 1.06 | **0.033** |
| *Care home* | 0.27 | 0.28 | 0.08, 1.10 | **0.047** |
| **Triage (CTAS) on index visit** |  |  |  |  |
| *5* | — | — | — |  |
| *4* | 3.37 | 3.59 | 1.27, 15.1 | **0.036** |
| *3* | 3.85 | 3.78 | 1.32, 16.0 | **0.030** |
| *2* | 6.75 | 7.14 | 1.80, 34.9 | **0.007** |
| **Time on stretcher at the ED** | 0.97 | 0.97 | 0.93, 1.00 | **0.035** |
|  | 1 OR = Odds Ratio, CI = Confidence interval for multivariate OR | | | |

C. Model characteristics from logistic regression model on 30-day revisits resulting in admission

| **Characteristic** | **Univariate OR***^1^* | **Multivariate OR***^1^* | **95% CI***^1^* | **p-value** |
| --- | --- | --- | --- | --- |
| **ZBI Score** | 1.02 | 1.02 | 0.99, 1.05 | 0.2 |
| **Charlson Score** | 1.18 | 1.17 | 1.06, 1.29 | **<0.001** |
| **Arrival method** |  |  |  |  |
| *Ambulance* | — | — | — |  |
| *Ambulant* | 1.46 | 1.57 | 1.01, 2.45 | **0.046** |
| **Caregiver annual income** |  |  |  |  |
| *No response* | — | — | — |  |
| *< 50,000$* | 0.74 | 0.75 | 0.46, 1.22 | 0.3 |
| *> or equal to 50,000$* | 0.37 | 0.36 | 0.18, 0.66 | **0.002** |
|  | ^1^OR = Odds Ratio, CI = Confidence interval for multivariate OR | | | |

### Additional File 6. Appendix F. Model output for sensitivity analyses

| **Characteristic** | **ZBI collected before revisit** | | | **ZBI collected after revisit** | | |
| --- | --- | --- | --- | --- | --- | --- |
|  | **OR***^1^* | **95% CI***^1^* | **p-value** | **OR***^1^* | **95% CI***^1^* | **p-value** |
| **ZBI Score** | 1.02 | 0.96, 1.07 | 0.4 | 1.02 | 0.97, 1.08 | 0.4 |
| **Previous ED visits** | 1.03 | 0.77, 1.29 | 0.8 | 1.05 | 0.89, 1.30 | 0.6 |
| **Covid-19 Period** |  |  |  |  |  |  |
| *Pre-pandemic* | — | — |  | — | — |  |
| *Wave 1* | 0.80 | 0.19, 2.84 | 0.7 | 0.43 | 0.09, 2.00 | 0.3 |
| *Between Wave 1 and Wave 2* | 3.93 | 0.79, 17.3 | **0.075** | 1.45 | 0.24, 13.3 | 0.7 |
| *Wave 2* | 1.13 | 0.35, 3.45 | 0.8 | 1.59 | 0.49, 5.45 | 0.4 |
| *Between Wave 2 and Wave 3* | 0.20 | 0.01, 1.77 | 0.2 | 0.39 | 0.01, 7.14 | 0.5 |
| *Wave 3* | 1.72 | 0.26, 8.32 | 0.5 | 1.29 | 0.25, 7.70 | 0.8 |
| **ZBI Score * Covid-19 Period** |  |  |  |  |  |  |
| *ZBI Score * Wave 1* | 1.09 | 0.99, 1.21 | **0.074** | 1.02 | 0.91, 1.16 | 0.7 |
| *ZBI Score **  *Between Wave 1 and Wave 2* | 0.82 | 0.59, 1.00 | 0.13 | 0.96 | 0.78, 1.22 | 0.7 |
| *ZBI Score * Wave 2* | 1.01 | 0.91, 1.10 | >0.9 | 1.00 | 0.90, 1.12 | >0.9 |
| *ZBI Score **  *Between Wave 2 and Wave 3* | 0.98 | 0.80, 1.15 | 0.9 | 1.24 | 0.94, 2.17 | 0.3 |
| *ZBI Score * Wave 3* | 0.91 | 0.68, 1.07 | 0.4 | 1.03 | 0.87, 1.26 | 0.7 |
| **Previous ED visits * Covid-19 Period** |  |  |  |  |  |  |
| *Previous ED visits * Wave 1* | 0.81 | 0.38, 1.42 | 0.5 | 1.44 | 0.91, 2.55 | 0.2 |
| *Previous ED visits **  *Between Wave 1 and Wave 2* | 1.44 | 0.70, 2.92 | 0.3 | 0.94 | 0.52, 1.88 | 0.8 |
| *Previous ED visits * Wave 2* | 1.03 | 0.67, 1.53 | 0.9 | 0.96 | 0.67, 1.43 | 0.8 |
| *Previous ED visits **  *Between Wave 2 and Wave 3* | 2.16 | 1.22, 4.85 | **0.029** | 1.03 | 0.46, 2.48 | >0.9 |
| *Previous ED visits * Wave 3* | 1.16 | 0.69, 1.81 | 0.5 | 0.79 | 0.49, 1.24 | 0.3 |
| *^1^* OR = Odds Ratio, CI = Confidence Interval | | | | | | |
